# Remote, unsupervised functional motor task evaluation in older adults across the United States using the MindCrowd electronic cohort

**DOI:** 10.1101/2021.05.17.21257333

**Authors:** Andrew Hooyman, Joshua S. Talboom, Matthew D. DeBoth, Lee Ryan, Matt Huentelman, Sydney Y. Schaefer

## Abstract

The COVID-19 pandemic has impacted the ability to evaluate motor function in older adults, as motor assessments typically require face-to-face interaction. This study tested whether motor function can be assessed at home. One hundred seventy-seven older adults nationwide (recruited through the MindCrowd electronic cohort) completed a brief functional upper-extremity assessment at home and unsupervised. Performance data were compared to data from an independent sample of community-dwelling older adults (N=250) assessed by an experimenter in-lab. The effect of age on performance was similar between the in-lab and at-home groups for both the dominant and non-dominant hand. Practice effects were also similar between the groups. Assessing upper-extremity motor function remotely is feasible and reliable in community-dwelling older adults. This test offers a practical solution in response to the COVID-19 pandemic and telehealth practice and other research involving remote or geographically isolated individuals.

## Introduction

Assessing motor function in older adults is essential, as it is affected by neurologic conditions like stroke (Lang et al., 2013), Mild Cognitive Impairment (MCI) (Jekel et al., 2015), Parkinson’s disease (Roalf et al., 2018) and Alzheimer’s disease (Kluger et al., 1997). However, most clinical motor assessments require face-to-face administration and specialized medical equipment (e.g., dynamometry) (Milne & Maule, 1984), and experimental measures typically use motion capture (Heath et al., 1999; Owings & Grabiner, 2004), robotics (Pearce et al., 2012), or other expensive technologies (e.g., transcranial magnetic stimulation, electromyography, or magnetic resonance imaging) (Ferber et al., 2002; Resnick, 2000; Schambra et al., 2015). Thus, these motor assessments are not feasible in remote contexts and are limited in re-test frequency or longitudinal evaluation due to cost and time constraints. Due to the COVID-19 pandemic, many medical practices and research methods have shifted to remote, internet-based approaches (Klil-Drori et al., 2021; Rowley et al., 2019; Thornton, 2020), and many older adults are unwilling or unable to engage in face-to-face, in-person research (Roe et al., 2021). However, the objective motor assessments currently available cannot be done remotely due to instrumentation or supervision requirements (i.e., a piece of equipment and a test administrator is needed). These limitations are problematic for evaluating motor function, tracking disease progression over time, and measuring the efficacy of an intervention as an outcome variable, particularly for older populations who have been encouraged to remain isolated due to COVID-19. Thus, there is an urgent need for objective motor assessments that are feasible and reliable for remote administration in people’s homes.

A simple, low-cost upper extremity motor task has been developed as a more accessible and affordable assessment (Schaefer et al., 2015). It is fabricated from household items, requires minimal technology (only a stopwatch or timing device), and can be assembled and mailed for <$10. Research on the face-to-face version of the task (administered by an experimenter) has shown that older adult task performance is associated with cognitive status (Hooyman et al., 2020), visuospatial memory (Lingo VanGilder et al., 2019), and a one-year decline in activities of daily living (Schaefer et al., 2020), yet is not subject to sex differences (Hooyman et al., 2020). It is also feasible for stroke (Schaefer et al., 2013), Parkinson’s disease (Paul et al., 2020), and MCI (Schaefer et al., 2020; Schaefer & Duff, 2017) populations. These features collectively make this task amenable to a remote, at-home setting, but it has not been validated for such use to date.

To test the reliability of our motor assessment as an unsupervised, at-home tool, we utilized the MindCrowd electronic cohort (Huentelman et al., 2020). MindCrowd is an internet-based platform focused on neuroaging research that has collected participant data via online surveys and a brief cognitive assessment, as well as remote genotyping via mail (Talboom et al., 2019). Thus, the purpose of this study was to leverage the MindCrowd infrastructure to validate a remote, unsupervised version of our motor assessment within an in-home setting. To do so, we utilized a subsample of MindCrowd participants over age 40, and compared their at-home, unsupervised performance to data from an existing in-lab, supervised sample.

## Methods

### Participants

All participants in this study had no previous history of mental illness, neurologic disease, or injury (i.e., stroke, history of seizures, concussion diagnosis, brain disease, or arthritis of the hands or upper limbs). All participants reported normal visual acuity and absence of any peripheral sensory or motor loss/pathology. Although MindCrowd itself has users worldwide, all participants in this study resided in the United States.

Six hundred seventy-nine participants were recruited to this study via e-mail, which provided an overview of the motor task and the option to agree to participate if interested. If the participant consented (via e-mail), a kit containing the motor task, along with instructions for administration and reporting data, was sent to the mailing address provided by the participant. As of January 2021, 241 kits had been mailed to consented participants, and 177 participants (mean age = 59.13 years +/- 9.18; 132 female) had completed the task and reported their data back to MindCrowd. Thus, ∼1/3 of contacted MindCrowd users were willing to participate, ∼75% of whom completed the assessment with their dominant and non-dominant hand once consented. In this cohort, hand dominance was self-reported. The WCG IRB Institutional Review Board approved this portion of the study.

The MindCrowd data were then compared to an independent sample that had been previously completed collected in-lab (Hooyman et al., 2020) (N = 250, mean age = 73.12 years +/- 8.22, female = 129). All participants in the in-lab cohort were assessed on their dominant and non-dominant hand and provided written informed consent before participation following the World Medical Association Declaration of Helsinki. The Arizona State University and Utah State University Institutional Review Boards approved this portion of the study. A subset of 106 participants from the in-lab cohort (mean age = 71.29 +/- 8.67, female = 72) also completed three more trials of the motor task with the non-dominant hand to evaluate a practice effect. Hand dominance in this cohort was assessed with the Edinburgh Handedness Inventory (Oldfield, 1971).

### Motor task

Details regarding the administration of the motor task have been published previously (Schaefer et al., 2015; Schaefer & Duff, 2015; Schaefer & Hengge, 2016) and are viewable on Open Science Framework (https://osf.io/mebcq/?view_only=ce5d894a2490462ebb87d60222462558). Importantly, this task has been related to measures of daily functioning in older adults (Schaefer et al., 2020), demonstrating its ecological validity. Briefly, task performance involved 15 repetitions of acquiring and transporting two kidney beans (∼0.5cm^3^) at a time with a standard plastic spoon from a plastic ‘home’ cup (9.5 cm in diameter and 5.8 cm in height) to one of three ‘target’ cups that were the same size as the home container. The target containers were secured radially around the home container at −40°, 0°, and 40° at 16 cm. At the start of the trial, thirty beans were placed into the home cup (15 repetitions x 2 beans/rep). To replicate the experimental set-up at home, individual kits were mailed that had 4 cups, 30 beans, a spoon, written instructions, and a paper ‘gameboard’ that provided a visual of where the home and target containers should be placed relative to their body, along with tape adhesive to adhere the gameboard to a table while seated. Figure 1 illustrates an assembled view of the task. Participants started by reaching to the left target cup, then returned to the central cup to acquire two more beans to transport to the middle target cup, then the right target cup, and then repeated this 3-cup sequence five times for a total of 15 reaches. The trial ended once the last two beans were deposited into the last cup. Performance was measured as the amount of time it took to complete all 15 reaches, i.e., ‘trial time.’ Four trials were completed with the non-dominant hand (to evaluate a practice effect), and one trial was performed with the dominant hand. MindCrowd participants either timed themselves (61%) or were timed by a partner (39%), while an experimenter timed the in-lab participants.

**Figure 1.**
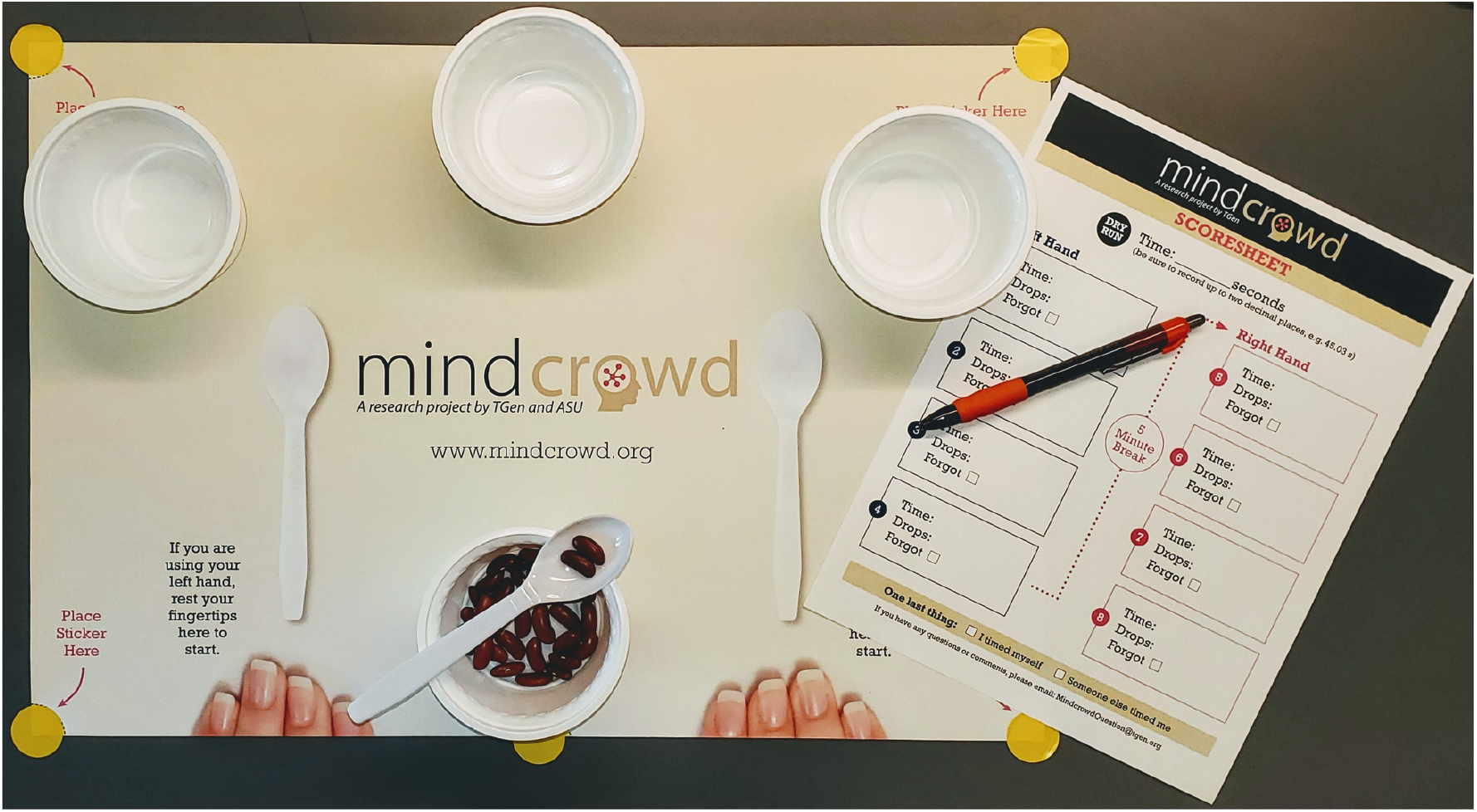
Example of assembled task kit. Written instructions and link to the video tutorial on task set-up were included.

### Statistical analyses

All analyses were performed in R (v4.0.0). To determine the reliability of the assessment, we performed a general linear model with task performance (i.e., trial time) as the dependent variable; group (MindCrowd vs. in-lab), sex, age, and hand were included as independent variables. Since the non-dominant hand performed four trials overall, only the first trial was used in this first analysis. To determine the similarity in practice effects of the non-dominant hand between MindCrowd and in-lab groups, we performed an autoregressive linear mixed-effects model with performance of the non-dominant hand across the four trials as the dependent variable, and trial number (1-4), group, age, and sex as fixed effects and random intercepts on the participant.

## Results

The general linear model showed no significant effect of group (MindCrowd vs. in-lab) on task performance (*p*=.2), indicating that data collected in-lab by an experimenter was comparable to data collected at home by the participant unsupervised. For example, the mean difference in task performance between in-lab and MindCrowd groups was only ∼2% (1.3 seconds). Regardless of group, there was a positive relationship between age and task performance (β_age_=.29, *p*<.0001, 95% CI=[.2, .38]), consistent with (Spedden et al., 2017). There was also an effect of hand dominance on task performance (β_dominantHand_=-8.62, *p*<.0001, 95% CI=[-9.46, −7.78]) (Fig. 2A), consistent with previous data showing that the dominant hand is faster (Schaefer, 2015). Furthermore, there was also no effect of sex on task performance (β_Sex_=1.21, *p*=.17, 95% CI=[-.51, 2.94]), again consistent with previous data (Hooyman et al., 2020). Within the MindCrowd cohort, there was no significant effect on task performance based on whether participants timed themselves or were timed by someone else (mean non-dominant hand difference = 2.6 seconds, 95% CI=[-1.7, 6.4], *p*=.18; mean dominant hand difference=.51 seconds, 95% CI=[-1.6, 2.7], *p*=.65). These results collectively show that age (and hand used) impact task performance, while the location and level of task supervision do not.

**Figure 2.**
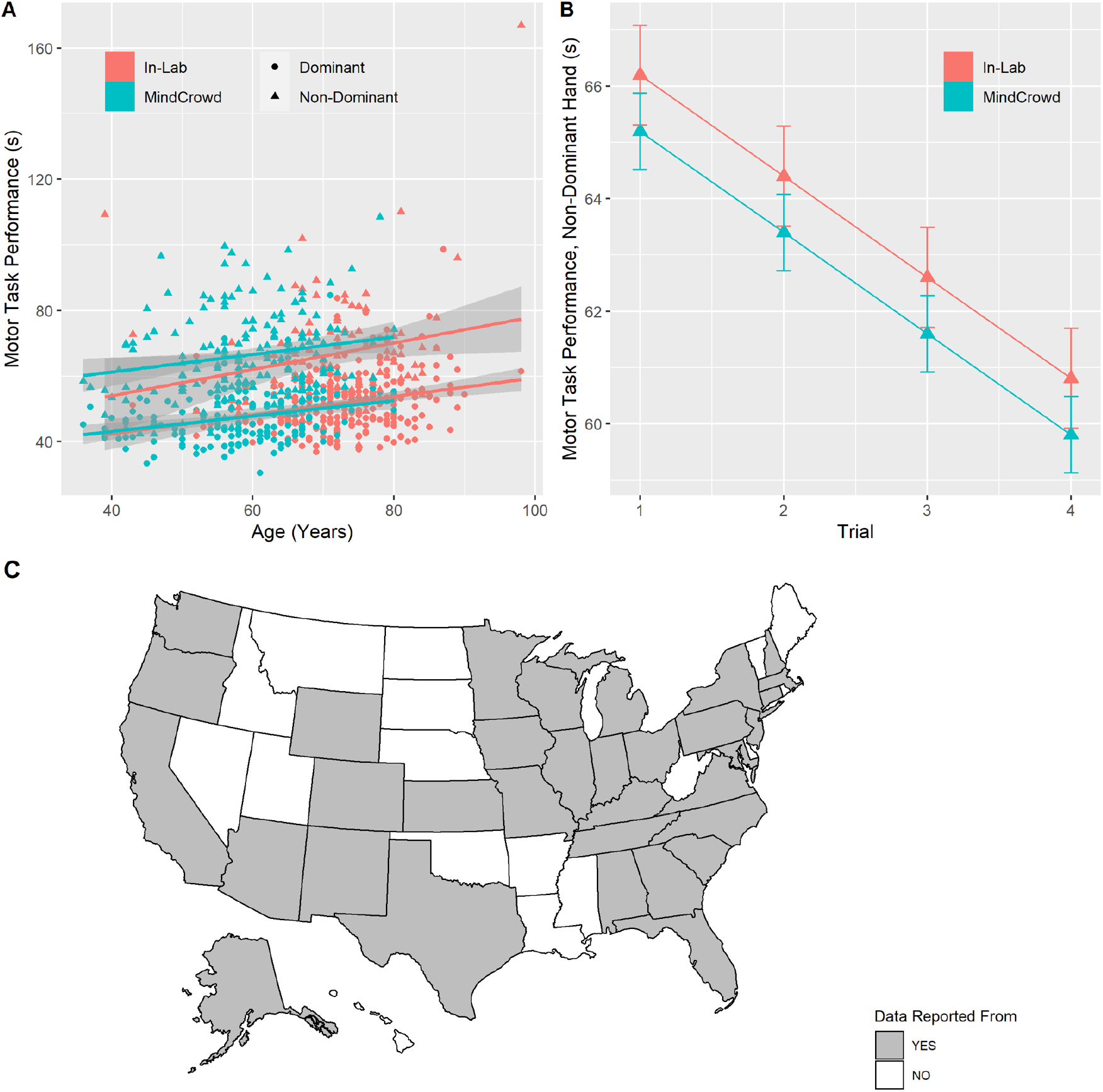
A) Relationship between age and motor task performance by group: in-lab (pink) vs. MindCrowd (blue); and hand: dominant (circle) vs. non-dominant (triangle). Gray shading indicates standard error. B) Estimated trial time of the non-dominant hand across four practice trials calculated from a linear mixed-effects model. In-lab (pink); MindCrowd (blue). Error bars = standard error. Note that the y-axis scale is different here than in panel A. C) US map with grey states showing where data were reported.

The autoregressive linear mixed-effects model also exhibited no effect of group on practice effects when the task is completed multiple times with the non-dominant hand (*p*=.12) (Fig. 2B). Consistent with earlier data, there was a significant effect of the trial (i.e., participants improved with practice) (β_trial_=-1.8, *p*<.0001, 95% CI= [-2.27, - 1.32]), but these results indicate that improvements in the motor task due to repeated exposure were also not dependent on location or level of supervision.

## Discussion

The purpose of this study was to validate a remote, unsupervised version of a functional motor assessment within an in-home setting. Results showed that motor performance collected in-home without supervision was not significantly different from data collected face-to-face in a laboratory setting. In other words, performance, and corresponding practice effects, measured in the home were not statistically different from those measured in the lab. This suggests that older adults nationwide can reliably perform this motor assessment remotely without supervision or clinical oversight.

The feasibility and reliability reported here demonstrate measurable benefits for preclinical research in older adults. First, once participants consented to participate and the kits were mailed, it took only 174 days for all 177 participants to perform the task and report their data. This rate is >1 participant a day, including weekends, whereas the rate of recruitment/participation in a face-to-face research study is often much slower, even before the COVID-19 pandemic. Second, the low cost and simplicity of the individual task components (e.g., beans, plastic spoon) allowed motor data to be collected from all over the United States. As shown in Figure 2C, data were collected from participants in 33 different states, a paradigm much different than what is feasible in other single-site studies that involve face-to-face assessments. The ability to collect across a geographically distributed sample can make research more inclusive, particularly for older adults who cannot drive or who do not have access to reliable public transportation (Park et al., 2010). Third, social isolation (due to the COVID-19 pandemic or otherwise) can substantially affect depression and psychological distress among older adults (Gorenko et al., 2021); thus, gerontological research must continue pursuing ways to engage and assess isolated older adults. Lastly, the feasibility and reliability of assessing motor function at home and unsupervised allows for clinical trial enrichment. Performance on the motor task used here has been linked to visuospatial processes (Lingo VanGilder et al., 2018, 2019), which have been shown to decline earlier than memory scores in cases of eventual Mild Cognitive Impairment (MCI) diagnosis (Schaefer & Duff, 2017). Furthermore, performance on this motor task improves the prediction of eventual functional decline in confirmed MCI (Schaefer et al., 2020). The remote version of this task would allow researchers or clinicians to monitor or screen individuals easily, regardless of geographical location, for study enrollment or neuropsychological follow-up.

A benefit of electronic cohorts is that they allow for more extensive data to be collected from more distributed samples. MindCrowd has collected a number of demographic, health, and lifestyle variables on its users (see Talboom et al., 2019), although very few of these have been included in the analyses presented here because of limitations in the in-lab sample. In fact, all shared data elements (e.g., age, sex) between the two cohorts were included in the analyses to consider as many covariates as possible. While this is a limitation of this study, this highlights the advantage of leveraging electronic cohorts for human subjects research. They allow for more extensive and distributed samples and enable a much more robust set of data to be collected than typical face-to-face laboratory research. With the remote version of this motor assessment validated, future studies can investigate how other factors, such as zip code, race/ethnicity, socioeconomic status, former occupation, marital status, co-morbidities, polypharmacology, and genetics affect motor function and motor decline in older adults.

To summarize, this study showed that this motor assessment could feasibly and reliably be collected remotely without supervision in older adults. We demonstrate that individuals across a range of geographical locations and ages were willing to participate in a study that involved the assembly, completion, and reporting of a task.

## Data Availability

Some de-identified data may be made available upon reasonable request.

## Acknowledgements

We would like to acknowledge Jessica Trevino, Alaina Dettmer, Britney Hill, and Shreyas Jejurkar for the assistance in assembling the kit components.

